# Development and evaluation of an online training program for palliative care in India

**DOI:** 10.1101/2024.08.26.24312585

**Authors:** Varun Raj Passi, Sreedevi Warrier, Rajalekshmi Balu, MM Sunilkumar, Parth Sharma

**Affiliations:** Association for Socially Applicable Research, Pune, India; Trivandrum Institute of Palliative Sciences, Pallium India, Kerala, India; Department of Community Medicine, Maulana Azad Medical College, Delhi, India

**Keywords:** Palliative Care, Pain Management, Distance Education, India, Terminal Care

## Abstract

**Objective:** Palliative care training at the undergraduate level is poor in India. With the need for palliative care rising in India and globally, it is possible to train physicians in resource-limited settings in palliative care via online training programs owing to ease of access and convenience. However, there is limited evidence available from India on the development and implementation of such a training program. This study aims to describe the development of an online training program offered by the Trivandrum Institute of Palliative Sciences (TIPS), Kerala, and the impact of the program on the confidence of physicians in managing various palliative care needs of their patients.

**Material and Methods:** The course was developed by an interdisciplinary expert team from TIPS. The course content was made keeping in mind the local sociocultural factors in India and was peer-reviewed by two external experts. The program was started in 2018 and updated and revised over the years. Currently, the program has 20 sessions, each lasting for 90 minutes. The course content was disseminated using project-ECHO’s tele mentoring model. To assess the impact of the training program, self-reported change in confidence from twenty-two batches of physicians, trained from January 2020 to August 2023. Feedback from participants was also assessed to identify areas of improvement in the training program.

**Results:** A total of 1159 physicians were trained during the study period. At the end of the course, 51.2% of the participants completed the evaluation survey and had a statistically significant (p<0.05) improvement in confidence in pain, gastrointestinal symptoms and breathlessness management, morphine prescription, and psychosocial communication. The duration of each session and the entire course was reported to be ideal by 88.6% and 87.9% of participants, respectively.

**Conclusion:** Our results show that online training can be effectively used to build confidence in physicians in managing various palliative care needs.

## 1. Introduction

India had an estimated 100.4 incident cancer cases per 100,000 people in 2022, which is expected to rise.^[1]^ The incidence of other non-communicable diseases like diabetes, hypertension, cardiovascular and respiratory diseases has also grown in India over the last few decades.^[2]^ As these chronic diseases eventually reach an end stage, their increasing burden necessitates a scaling up of palliative care services to address disease-related suffering.

In 2023, the need for palliative care in India was estimated to be 6.21 of every 1000 individuals which roughly translates to 8.4 million people.^[3]^ However, despite the increasing burden of diseases, access to palliative care remains poor. It was estimated that 98.39% of people living with end-stage cancer in India have their palliative care needs unmet.^[4]^ The Economist’s “Quality of Death” index ranked India at the 67th position out of the 80 countries that were studied.^[5]^ Palliative care services in India are also unequally distributed. While most of India’s population lives in rural areas, palliative care services are provided via tertiary and secondary centres clustered in large cities.^[6]^ This gap is more pronounced in the northern part of India.^[7]^

A major contributing factor to these is the lack of training of healthcare workers in palliative care in India. ^[8,9,10]^ Palliative care training can be divided into specialist education, generalist education, and palliative education approach.^[11]^ Specialist education offers in-depth training solely focused on a specialty, allowing providers to address complex palliative care needs. Specialist training in palliative medicine was recognized by the erstwhile Medical Council of India in 2010^[12]^ and as of June 2023 is available as a three-year course in 17 teaching hospitals.^[13]^

Generalist education provides shorter courses enabling specialists or general practitioners to offer palliative care alongside their primary practice, mainly for patients with less complex needs. The palliative education approach emphasises that all healthcare providers dealing with chronic and life-limiting conditions should possess basic palliative care skills, understand its principles, and integrate them into practice. The Competency Based Medical Education (CBME) states in its objectives that an Indian Medical Graduate should be able to understand and provide palliative care to their patients.^[14]^ The MBBS curriculum was updated in 2019 to include modules on palliative care. However, there remains a need for trained educators to teach palliative medicine to medical students.^[15]^

Palliative care training courses can equip Indian doctors with the skills needed to provide effective palliative care services. Usage of the online medium allows disseminating the educational content to a large audience, making it more accessible.^[11]^ While such courses could help improve basic awareness amongst healthcare providers, they need to be designed keeping the local sociocultural context and patient needs in mind. Additionally, their efficacy needs to be studied.

Our study aimed to describe the development of an online training program in palliative care for physicians-the Foundation Course in Palliative Medicine (FCPM), offered by the Trivandrum Institute of Palliative Sciences (TIPS). We also aimed to study the impact of the program on the confidence of participants in managing various palliative care needs.

## 2. Material and Methods

### 2.1 Program Background

Since 2008, TIPS has offered an in-person 6-week course in palliative medicine-The Certificate Course in Pain and Palliative Medicine (CCPPM). Experience from conducting the CCPPM inspired the creation of a virtual training program to disseminate palliative care training to a wider audience.

In January 2017, TIPS launched a virtual “Grand Rounds Program” (GRP) in collaboration with Project Extension of Community Health Outcomes (ECHO) to discuss the history and management of patients requiring palliative care. Project ECHO is an online technology-enabled capacity-building model, which aims to improve the knowledge and skills of community-level healthcare providers via teaching and mentorship. This is achieved by multipoint video conferencing which connects local healthcare providers with specialists at a hub site. The sessions are held regularly and follow a structured format of didactic teaching and case presentations.

The GRP consisted of 15 palliative care doctors. The sessions were 2-3 hours long and were held every 2 weeks. Each session involved 4-5 case discussions along with a 15-minute didactic session on a pre-decided topic. The cases were brought in before the session from any of the interconnected centres. Feedback from the attendees of this program informed the creation of the FCPM to disseminate palliative care education to doctors with no background training in palliative care.

### 2.2 Expert Team Development

An interdisciplinary expert team was established to develop the FCPM. This included five healthcare workers (two doctors from TIPS, two doctors who were recruited from the grand rounds program, and one nurse) with expertise in palliative care along with two technical support specialists and one program coordinator. During the planning phase of the course, the expert team met for multiple sessions to develop the learning program in concordance with the palliative care needs of the country. Since the launch of the first batch in June 2018, the expert team met for debriefing sessions after each batch was conducted to modify the course structure and content based on the feedback received from the participants.

### 2.3 Course Structure and Content

The course was designed specifically keeping in mind the learning needs of doctors with no background training in palliative care. The FCPM could be taken by any doctor with an MBBS or BDS degree (or equivalent) with a valid registration in their country.

The structure and content of the course were decided by discussion amongst members of the expert team during the planning phase of the course. The majority of topics covered were adapted from the CCPPM, and modified to meet the requirements of an online format.

The course was designed to have 20 sessions with each session being 1.5 hours long. The sessions were conducted in English. Each session was structured in the following manner:

First 5 minutes: General introduction of the faculty and participants

Next 40 minutes: Presentation by the faculty

Next 15 minutes: Discussion on the presentation

Next 5 minutes: A presentation by one of the participants on a patient’s health condition

Next 25 minutes: Discussion based on the patient’s story

The topics covered in the FCPM included:

1. Introduction to palliative care
2. Introduction to pain and its management
3. Medical ethics
4. Communication
5. Palliative care in non-cancer illness
6. Gastrointestinal and Respiratory symptom management
7. Wound management
8. Psychosocial issues
9. Delirium
10. Palliative care emergencies
11. Organising of palliative care services
12. End-of-life care
13. Narcotic Drugs and Psychotropic Substances (NDPS) Act and opioid availability

The course content and structure were peer-reviewed by two palliative care experts outside of the expert team. The course was finally launched in June 2018

### 2.4 Learning Materials

Video recordings of the live sessions were shared with the participants after each session along with other presentation materials (such as slides and key learning points). In addition to these, research articles pertaining to the topics discussed were shared as well. The participants were encouraged to use two textbooks as references: *Introducing Palliative Care* ^[16]^ and *An Indian Primer of Palliative Care.*^[17]^

An online group with all the participants was created on a chat platform. This group was moderated by faculty. The intention for this was to keep the participants engaged and create a sense of community. Participants were encouraged to ask questions in the group chat and share additional learning resources.

### 2.5 Method of Program Evaluation

At the time of registration, a demographic profile of all the participants was collected which included age, gender, qualification, healthcare sector they work in, country of residence, and state of residence.

At the end of the course, participants were asked to fill out an evaluation survey. The survey collected demographic data including whether they were actively practising medicine at present, and if they were currently seeing any palliative care patients. The survey also assessed the effect of the course on the participant’s confidence in managing various palliative care needs. This was done via a retrospective pretest-posttest design, wherein the participants were asked at the end of the course to rate their confidence before and after the course on a scale of 1 to 10, with 1 indicating “not confident at all” and 10 indicating “very confident”.

Feedback was also collected regarding the duration and timing of the sessions and the total duration of the course.

### 2.6 Data Analysis

The data was analysed using the programming language R. Quantitative variables were expressed as mean ± standard deviation, and qualitative variables were expressed as absolute and relative frequencies (percentages). Paired-t test was used to evaluate differences in confidence in managing various palliative care needs before and after the course. A p-value of less than 0.05 was considered to be significant.

### 2.7 Ethical Approval

The study was exempted from ethical review by the Internal Ethics Committee of Trivandrum Institute of Palliative Sciences on 8th July 2024. The ethics approval number of the study is TIPS/IEC-1/2024Exm.

## 3. Results

The study period was from January 2020 to August 2023. During this period, 22 batches of the FCPM were conducted.

### 3.1 Demographic characteristics

A total of 1159 physicians registered for the course during the study period. Out of these, 594 (51.2%) completed the evaluation survey.

Demographic characteristics of participants who registered for the course are in **Table 1** and those who completed the evaluation survey are in **Table 2**.

**Table 1:**
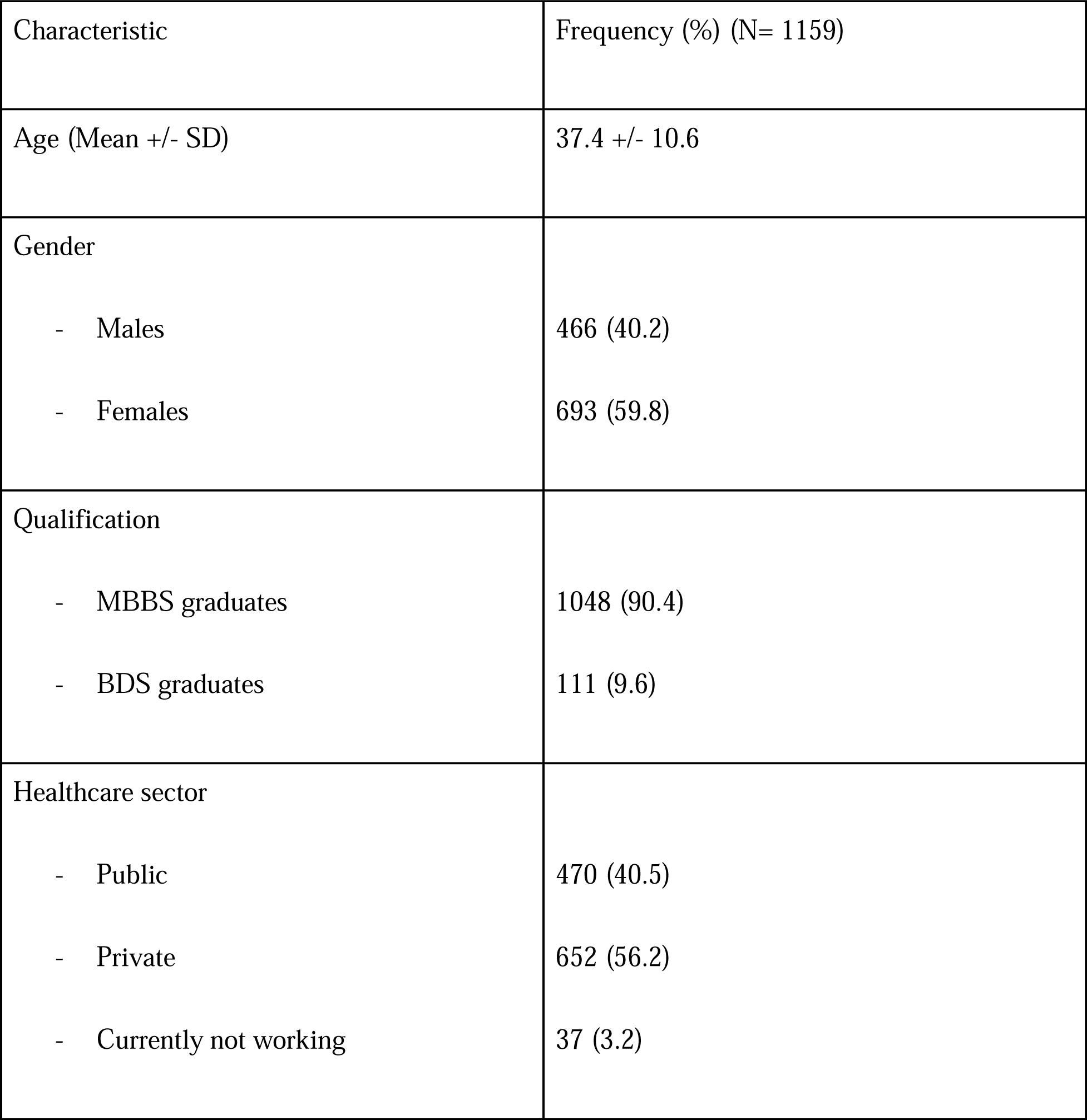
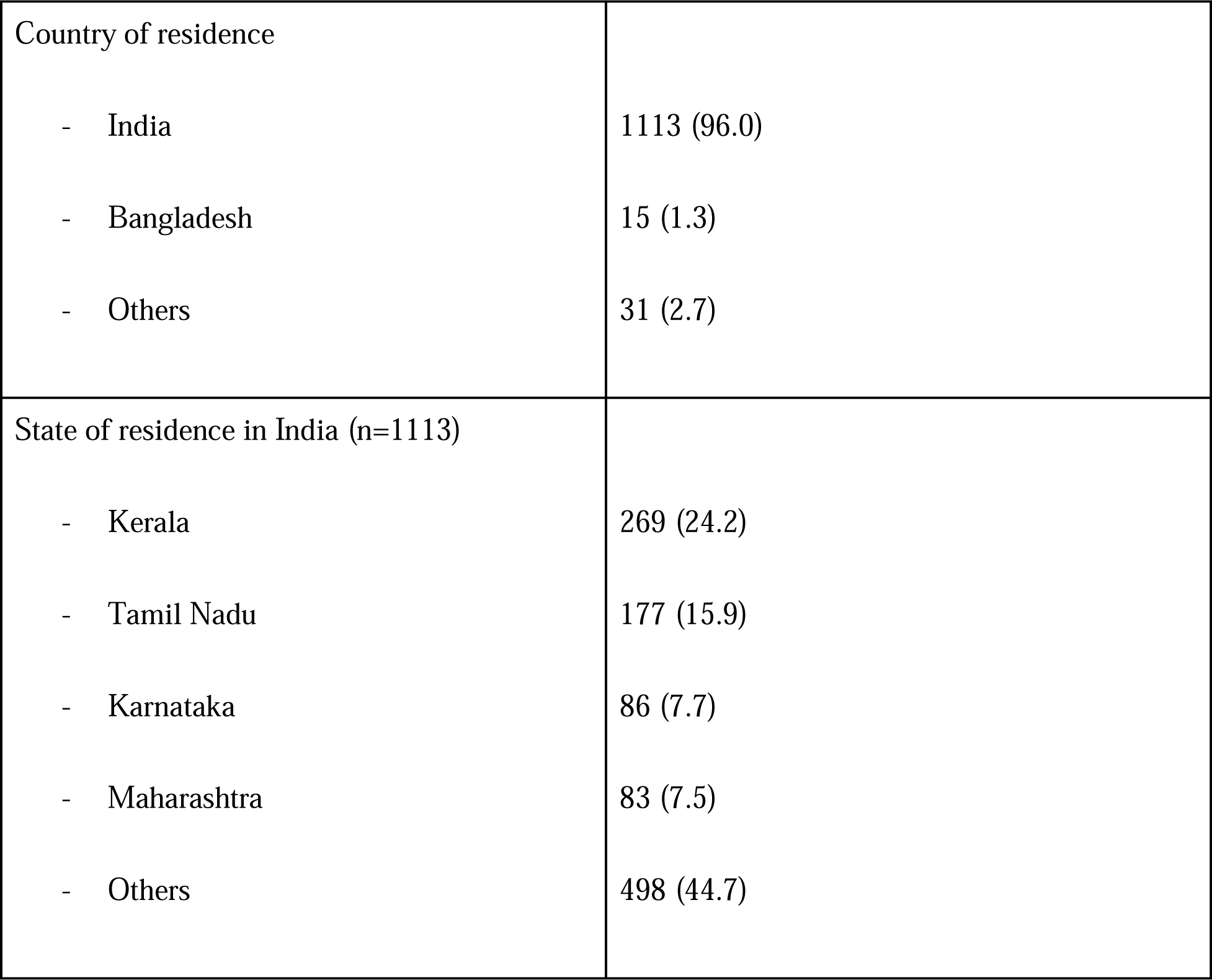
Demographic characteristics of registered participants.

**Table 2:** Demographic characteristics of participants who completed the evaluation survey.

| Characteristic | Frequency (%) (N=594 ) |
| --- | --- |
| Age (Mean +/- SD) | 38.7 +/- 10.7 |
| Gender |  |
| - Males | 227 (38.2) |
| - Females | 367 (61.8) |
| Qualification |  |
| - MBBS graduates | 538 (90.6) |
| - BDS graduates | 56 (9.4) |
| Healthcare sector |  |
| - Public | 216 (36.4) |
| - Private | 353 (59.4) |
| - Currently not working | 25 (4.2) |
| Country of residence |  |
| - India | 565 (95.1) |
| - Others | 29 (4.9) |
| State of residence in India (n=565) |  |
| - Kerala | 123 (21.8) |
| - Tamil Nadu | 58 (10.3) |
| - Karnataka | 51 (9.0) |
| - Maharashtra | 47 (8.3) |
| - Others | 286 (50.6) |
| Currently practicing medicine |  |
| - Yes | 497 (83.7) |
| - No | 97 (16.3) |
| Currently seeing palliative care patients |  |
| - No | 273 (46.0) |
| - Yes, but do not work in the palliative care department | 239 (40.2) |
| - Yes, I work in the palliative care department | 82 (13.8) |

### 3.2 Impact of online training on confidence in managing palliative care needs

The evaluation survey showed a statistically significant improvement in confidence in managing all the palliative care needs that were assessed (**Table 3**). These include confidence in managing pain, confidence in prescribing oral and IV morphine, confidence in managing GI symptoms, confidence in managing breathlessness, and confidence in discussing psychosocial issues with patients.

**Table 3:** Pre-test and post-test confidence scores of participants.

|  | Pre-test mean score<br>(N=594) | Post-test mean score<br>(N=594) | p-value |
| --- | --- | --- | --- |
| Confidence in pain management | 5.19 +/- 2.02 | 8.44 +/- 1.18 | <0.001 |
| Confidence in prescribing oral or IV morphine | 3.90 +/- 2.71 | 8.10 +/- 1.69 | <0.001 |
| Confidence in managing GI symptoms | 5.34 +/- 2.25 | 8.55 +/- 1.30 | <0.001 |
| Confidence in managing breathlessness | 4.80 +/- 2.39 | 8.29 +/- 1.40 | <0.001 |
| Confidence in<br>discussing<br>psychosocial issues | 4.90 +/- 2.32 | 8.55 +/-1.20 | <0.001 |

### 3.3 Feedback from participants

A majority of the participants (477 (80.3%)) who completed the evaluation survey were comfortable with the present timing of sessions between 5 to 6:30 pm. While 301 (50.6%) considered it ideal, 156 (26.3%) and 83 (14.0%) of the participants preferred the timing of the sessions to be between 3 to 4:30 pm and 6 pm to 7:30 pm, respectively. 522 (87.9%) of the participants considered the total number of sessions (20 sessions) in the course to be ideal, and 526 (88.6%) considered the duration of each session (90 minutes) to be ideal. **Figure 1** shows the ideal time of the course for the participants.

**Figure 1.**
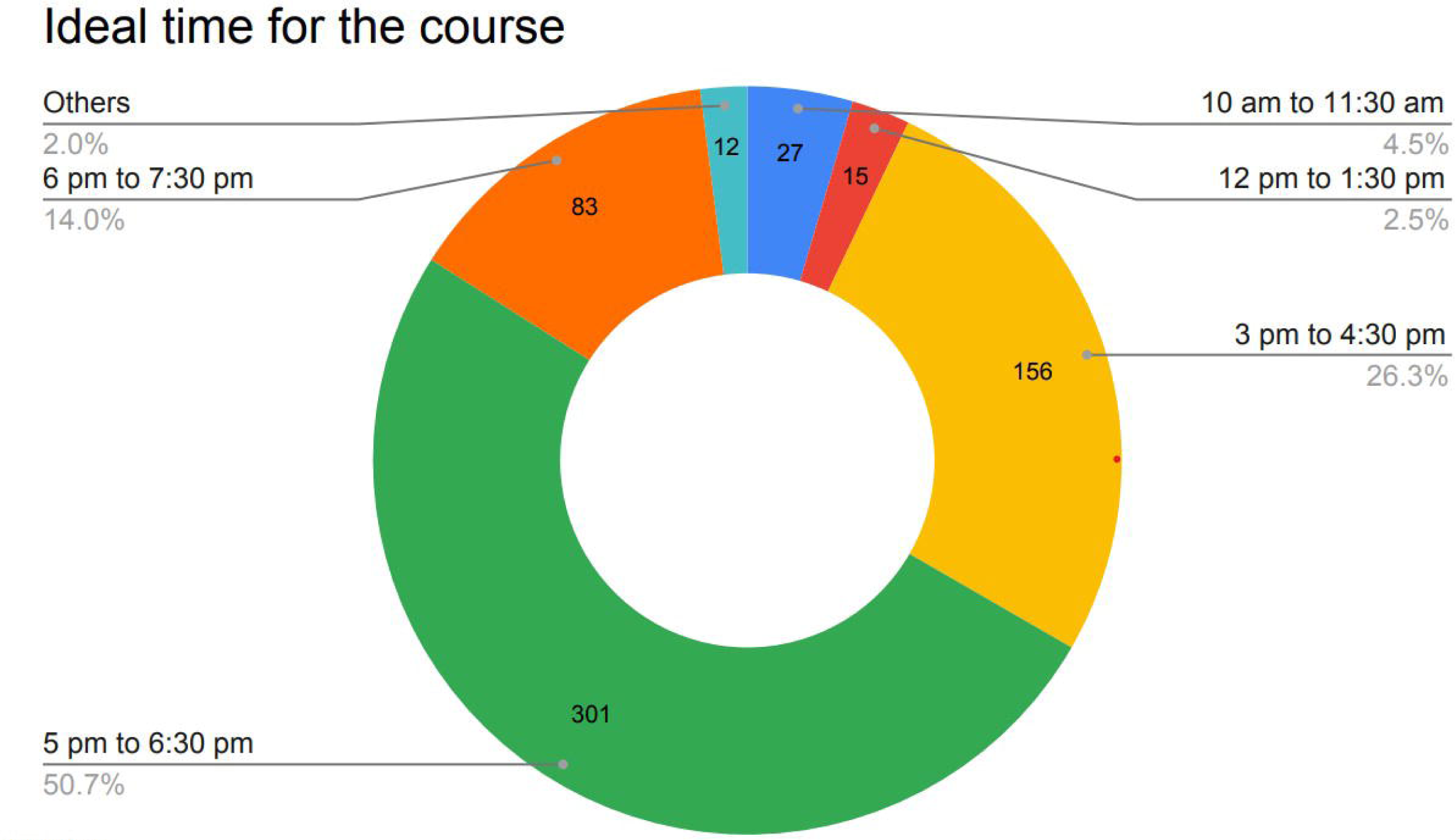

## Discussion

We have described our experience in developing, implementing, and evaluating a virtual training program in palliative care for doctors. The program’s structure and content were developed and validated by palliative care experts from India and continuously modified based on feedback from each batch. A survey assessing the course’s impact on the confidence of the participants in managing various palliative care needs was administered at the end of the program. A retrospective pretest-posttest design was used to make the survey which allowed the respondents to gauge prior levels of confidence compared to current levels after the course by providing a consistent frame of reference.^[18]^ The majority of the participants in our program were residents of India (96%) and held an MBBS degree (90.4%). 45.9% of the participants were from three states in the southern part of India-Kerala, Tamil Nadu, and Karnataka where the general availability of palliative care services is better.^[7]^ There were no significant differences in the demographic characteristics between those who registered for the course and those who completed the evaluation survey.

In resource-limited settings, Project ECHO has emerged as an innovative strategy to disseminate medical education. While there have been several studies that have described using Project ECHO to provide medical education to healthcare workers, a 2019 systematic review found very few studies from LMICs and also found most of them to be of poor quality ^[19]^. Since then, a few studies from India, Bangladesh, and Nepal have described virtual palliative care education programs using Project ECHO. ^[20, 21, 22, 23]^

Our course incorporated previously described modifications for LMICs to the original project ECHO model.^[22]^ These include the involvement of key local stakeholders in the leadership team, and continuous modification of the course by incorporating feedback from participants. We also encouraged asynchronous learning by creating a faculty-monitored social media group and providing the participants with video recordings of the sessions and presentation materials. Such modifications can facilitate learning, especially in resource-limited settings, and should be considered when implementing virtual training programs in LMICs. The involvement of local palliative care experts in the leadership team helped tailor the course content to the regional context. An example of this is the inclusion of “The Narcotic Drugs and Psychotropic Substances (NDPS) Act and opioid availability” in the course, which is specifically relevant to the Indian context where access to opioids is a major barrier to adequate pain control.^[24]^

The results of our evaluation survey showed that the participants showed a significant improvement in their confidence in managing all the palliative care needs that were tested. These results were concordant with studies that have evaluated the impact of palliative care training programs incorporating Project ECHO. These include studies from high-income countries (HICs) like Canada,^[25]^ and LMICs in Asia like Nepal, Bangladesh, and India ^[20,22,23]^ and Africa ^[26]^. While these programs had individual differences in their structure and course, they show the utility of Project ECHO in disseminating palliative education to healthcare workers across varying backgrounds. Our study was unique in that it solely focussed on training physicians. Further, unlike other studies from South Asia which primarily focused on paediatric palliative care, our study focussed on training in general palliative care.

Our results show that online training can be used to increase the confidence of doctors in managing the palliative care needs of their patients. Project ECHO’s unique tele mentoring model offers a feasible way to spread medical education to a large number of participants. This is especially useful in resource-limited settings where arranging in-person training is difficult. Given the poor quality of undergraduate training in palliative care, such courses can be used to build palliative care capacity in the country. Therefore, organisations such as the National Medical Council (NMC) should consider recognizing and implementing distance learning courses like ours at the national level to improve access to palliative care in the country.

### Study limitations and strengths

Our study had a few limitations. First, only 51.2% of the eligible participants completed the end-evaluation survey, which could lead to selection bias favouring participants who found the course to be more engaging and useful. Second, participants were asked to self-report their confidence in managing palliative care needs which may not represent their actual level of learning and capabilities. Third, we were unable to assess the patient-level impact of our course. Future research should assess the effect of these courses on the quality of care received by patients.

Despite these limitations, our study has many strengths. We developed and evaluated an innovative virtual palliative care training program for physicians. Our course was consistently modified based on feedback, keeping in mind the learning needs of our participants. The results of our study show the utility of online education in disseminating palliative care training to doctors. Our study also had a relatively larger sample size of 594 physicians who completed the evaluation survey, compared to other recent similar studies from South Asia. These include studies from India, Nepal, and Bangladesh where 51, 27, and 18 participants completed the evaluation survey respectively. ^[20,23,24]^

## Conclusion

Our study shows that virtual training is an innovative strategy to improve confidence in doctors in managing various palliative care needs. Therefore, online training programs have the potential to bridge gaps in access to palliative care by increasing trained palliative care providers, especially in resource-limited settings. Future research should assess the impact of these courses on patients and their families.

## Data Availability

All data produced in the present study are available upon reasonable request to the authors

## Notes

### Competing Interest Statement

Parth Sharma is the founding editor of Nivarana.org, a public health information and advocacy platform and has received honorarium for articles published in The Harvard Public Health Magazine, Think Global Health, The Hindu and The Wire.

### Funding Statement

This study did not receive any funding.

### Author Declarations

Ethics Committee of Trivandrum Institute of Palliative Sciences waived ethical approval for this work.

